# Survival of health workers infected by SARS-CoV-2 in the context of vaccination against COVID-19 in Peru

**DOI:** 10.1101/2021.08.03.21260614

**Authors:** Stefan Escobar-Agreda, Javier Silva-Valencia, Leonardo Rojas-Mezarina, Javier Vargas-Herrera

## Abstract

**Objectives:** To evaluate the survival of health workers infected by SARS-CoV-2 in the context of the vaccination process against COVID-19 in Peru.

**Methods:** A survival analysis was performed using data from national health databases. Data from people between 18 and 59 years old infected with SARS-CoV-2 as evidenced by molecular or antigenic tests were included. Kaplan Meier graphs were produced to compare the survival of health workers and the rest of the population during 2021 and health workers during the first and second wave of mortality in Peru in 2020 and 2021, respectively.

**Results:** Data from 998,295 people were included. The average age was 41.2 years (SD 15.8) and 485,167 (48.6%) were women. A higher level of survival of health workers after vaccination was found compared to the general population and to the population of health workers before vaccination. It was evidenced that, at the beginning of the second wave, the risk of dying for health workers was twice that of the first wave (HR = 2). After vaccination (in the sixth month of the second wave), the risk of dying decreased to 87.5% less than in the first wave (HR = 0.125).

**Conclusions:** A positive change has been evidenced in the level of survival of health workers infected by SARS-CoV-2 during the context of vaccination against COVID-19 in Peru.

## INTRODUCTION

COVID-19, a disease caused by the SARS-CoV-2 virus, has generated a significant impact on the health of the world population not only due to its ability to spread but also due to the risk of causing death in a sector of the population (1). Peru currently constitutes one of the countries with the highest mortality rate from this disease worldwide (2). In this context, the Peruvian government began coordination for the implementation of vaccination against COVID-19 in our country since the beginning of 2021 (3), given its recognition as one of the main resources to combat the pandemic at the population level (4). The scheme of the Vaccination Plan against COVID-19 in Peru, prioritized the vaccination of health workers as it is one of the populations most exposed to this virus (3), which to date has suffered a significant number of deaths in our country.

Some reports at the national level have preliminarily evaluated the potential effect of the vaccination process on health workers, showing a reduction in hospitalization and mortality rates of medical personnel during the first months of the vaccination period in Peru (5). However, to date it is not yet known whether there has been any variation in the level of survival of health workers infected by SARS-CoV-2, during the vaccination period compared to last year or with respect to the rest of the year. adult population, which, up to the moment of the present analysis, had not yet been vaccinated, according to the proposed national scheme. Additionally, it is important to know if these comparisons vary according to sex, given the evidence that indicates the difference in risk of mortality from COVID-19 between these subgroups (6).

In Peru, there are various national health information systems that include nominal information on people who underwent laboratory tests for the diagnosis of COVID-19 (NETLAB and SICOVID), health care and administrative personnel (INFORHUS) and deceased persons (SINADEF). Based on this information, this study aims to evaluate the survival of health workers infected by SARS-CoV-2 during the context of the vaccination process against COVID19 in Peru.

## METHODS

A secondary data analysis was performed using the databases of SINADEF (registry of deceased), INFORHUS (registry of health workers), SICOVID (registry of antigen tests for the diagnosis of SARS-Cov 2) and NETLAB (registry of molecular tests for the molecular diagnosis of SARS-Cov 2); which contain information about the people examined to determine their infection status by the SARS-CoV-2 virus through molecular tests, antigenic tests, their occupation to be classified as health workers and deceased persons at the national level respectively.

Records of persons between 18 and 59 years of age corresponding to their last positive test for SARS-CoV-2 as evidenced by molecular test (MT) or antigenic test (AT) were included. Records that did not have identity document data were withdrawn, or when the date of obtaining the sample was after the date of death, or whose date of death was after 60 days from the date of collection of the sample (for molecular or antigenic tests). To obtain estimates at the national level, no sampling was performed and all observations that met the selection criteria were included.

Based on these criteria, two study subpopulations were defined: people with a positive test for SARS-CoV-2 by MT or AT in Peru in 2021; and health workers with a positive test for SARS-CoV-2 by MT during the first wave and health workers with a positive test for MT or by AT, during the second wave of COVID-19 in Peru.

### Result variable

#### Decease

It was classified as “Deceased” if, according to its identity document, it is registered in the SINADEF database of deaths, and the rest were classified as “Not deceased.” In addition, data on the date of death were included to determine survival time.

### Exposure variables

#### Health personnel

All those people who, according to their identity document, are registered in the INFORHUS database updated until January 2021, were classified as “health workers”. To optimize this classification, it was added also as health workers to those people who were not registered in INFORHUS, but who have self-reported that they have an occupation as health workers: “doctor”, “nurse”, “obstetrician”, etc., at the time of obtaining of the sample. <u>Wave of COVID-19</u>. People included in the study were classified as either SARS-CoV-2 infected during the first wave, or as infected during the second wave of COVID-19 transmission. The start of each wave was defined as the first day of the epidemiological week in which there was an approximate 75% increase in deaths with respect to the historical average of deaths according to SINADEF data. Given the availability of data until June 30, 2021, an analysis time of 182 days was considered for each wave (∼ 6 months).

### Covariates

The following covariates were included: sex, classified as “male” or “female”; age, in years; insurance, “have health insurance” or “do not have health insurance” and “belong to the Lima region” or “do not belong to the Lima region”.

Statistical analyzes were performed in the Stata v16.0 statistical package for Windows. Initially, a descriptive analysis was carried out to evaluate the characteristics of the health workers and the general population infected by SARS-CoV-2 included in the study. For the survival analysis, Kaplan Meier curves were elaborated to compare the survival of health workers and the general population infected by SARS-CoV-2 during the year 2021 and the survival of health workers in the first and second wave during the COVID-19 pandemic in Peru in 2020 and 2021. For this last comparison, risk ratios were estimated (Hazard Ratios, HR) and risk differences (Attributable Risk Reduction, ARR) to mortality between both waves, indicating their 95% confidence intervals. For this, Cox regressions and polynomial models were used to graph the evolution of these indicators during the period of study for both waves. To improve the visualization of the survival of the populations studied in the study period, some important milestones were defined corresponding to the date of the start of vaccination (February 9, 2021), start of the second dose application (March 1, 2021), and start of the protection period (14 days after the start of the second dose). This last period was defined based on the time of protection suggested for the Sinopharm vaccine used for the vaccination of health workers in Peru.

The present study was based on a secondary analysis of data contained in the databases of the routine health information systems of the National Institute of Health and the Ministry of Health, which are shared mutually. The access and use of the information to these databases was authorized by the owners of the databases and was carried out as part of the competences of the National Institute of Health, within the framework of public health surveillance and in the context of the attention of the COVID19 pandemic in Peru. All the information used in the preparation of the analyzes presented has been kept strictly confidential by the researchers solely for the purposes of this study.

## RESULTS

998,295 people between 18 and 59 years old, whose last molecular or antigenic test was positive for SARS CoV 2 since the beginning of the pandemic, were studied. The mean age was 41.2 years (SD: 15.8), 485,167 (48.6%) were women, 466,459 (46.7%) came from Lima, 34,424 (3.5%) were health workers, and 35,981 (3.6%) had died according to their registration in SINADEF. Table 1 shows the characteristics between the health workers and the rest of the population included in the study.

**Table 1.** Characteristics of health workers and the general population, positives for COVID-19 by molecular or antigenic test in Peru, 2021. (N = 998>,295)

| **Características** | **Health worker,**<br>**n (%)**<br>**N=34,424** | **No Health worker,**<br>**n (%)**<br>**N= 963,871** |
| --- | --- | --- |
| **Age (years)** | 40.0 ± 10.9 | 41.3 ± 16.0 |
| **Sex** |  |  |
| Female | 22502 (65.4) | 462665 (48.0) |
| Male | 11922 (34.6) | 501206 (52.0) |
| **Provenance** |  |  |
| No Lima | 13616 (39.6) | 452843 (47.0) |
| Lima | 20808 (60.5) | 511028 (53.0) |
| **Death\*** |  |  |
| No death | 34071 (99.0) | 928243 (96.3) |
| Death | 353 (01.0) | 35628 (03.7) |

\* SINADEF database

### Survival of health workers compared to the general population

Since the beginning of the second wave, the Kaplan Meier curves showed a greater survival of health workers infected with the SARS CoV 2 virus compared to the rest of the population positive for SARS-CoV-2 in Peru. This difference was much more pronounced, due to a flattening of survival in health workers from March 15, 2021 (day 374 of the pandemic), date corresponding to 14 days after the start of the application of the second dose. (See graph 1).

**Graph 1.**
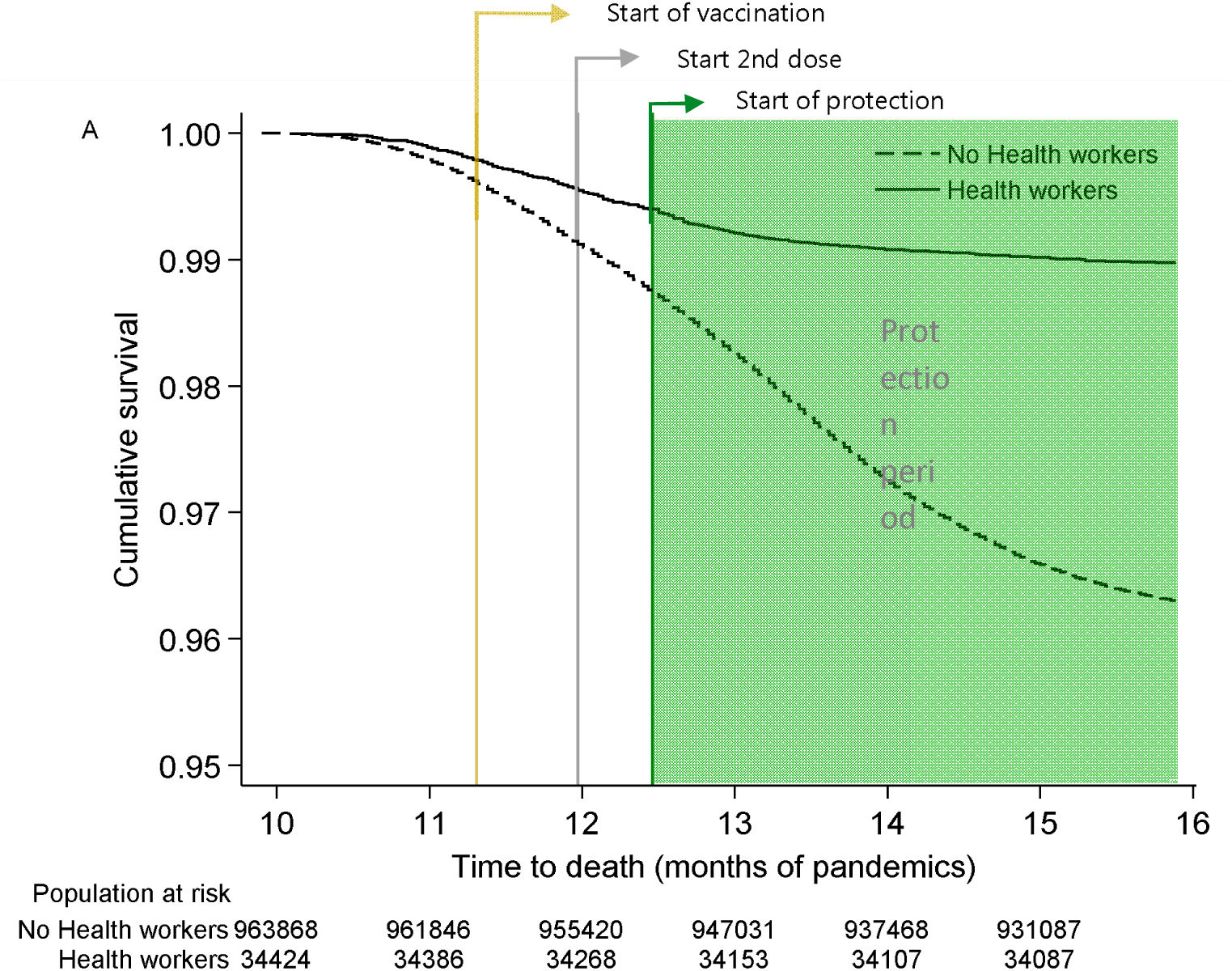
Survival levels (Kaplan Meier) of health workers (RHUS) and the rest of the population infected by SARS-CoV-2 (molecular test or antigenic test), during the COVID-19 pandemic in Peru, 2021 (N = 998,292).

### Survival of health workers in the first and second waves

At the beginning of the second wave, the mortality of health workers was higher compared to the first wave. However, later, the survival curve during the second wave shows a flattening, specifically from the beginning of the vaccine protection period, two weeks after the moment in which the application of the second dose began (March 15), reducing the gap of survival between the two waves. (see graph 2).

**Graph 2.**
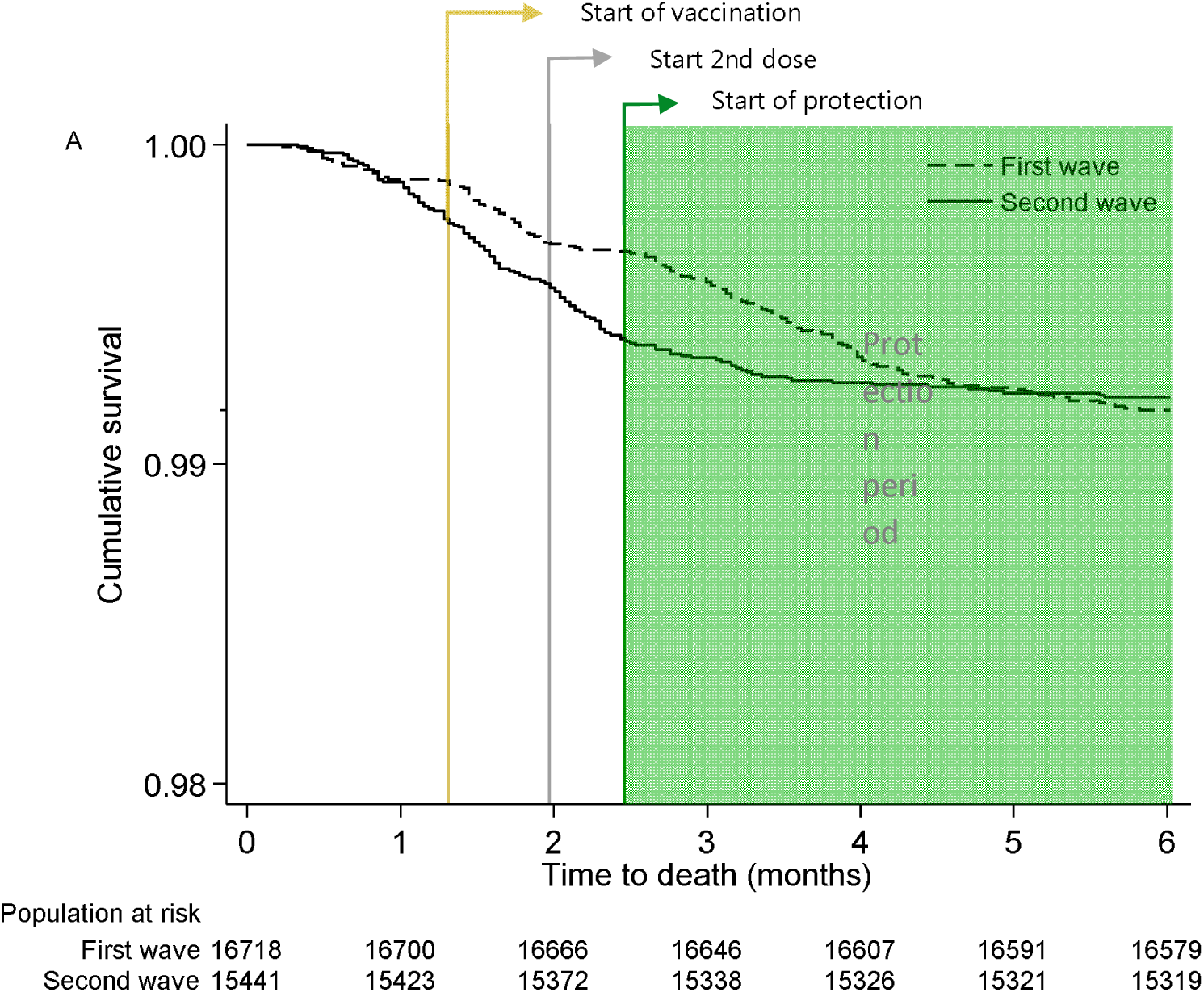
Survival levels (Kaplan Meier) of health workers infected by SARS-CoV-2, during the first and second wave of COVID-19 in Peru, 2020-2021. (N = 32,159)

Specifically, at the time the vaccination began, a positive difference in the risk of mortality of health workers was observed during the second wave with respect to the first (risk difference> 0), which was reduced to until it is canceled towards the end of the study period (risk difference ∼ 0). The absolute risk reduction up to the sixth month of the second wave was less than zero compared to the first wave. (See graph 3)

**Graph 3.**
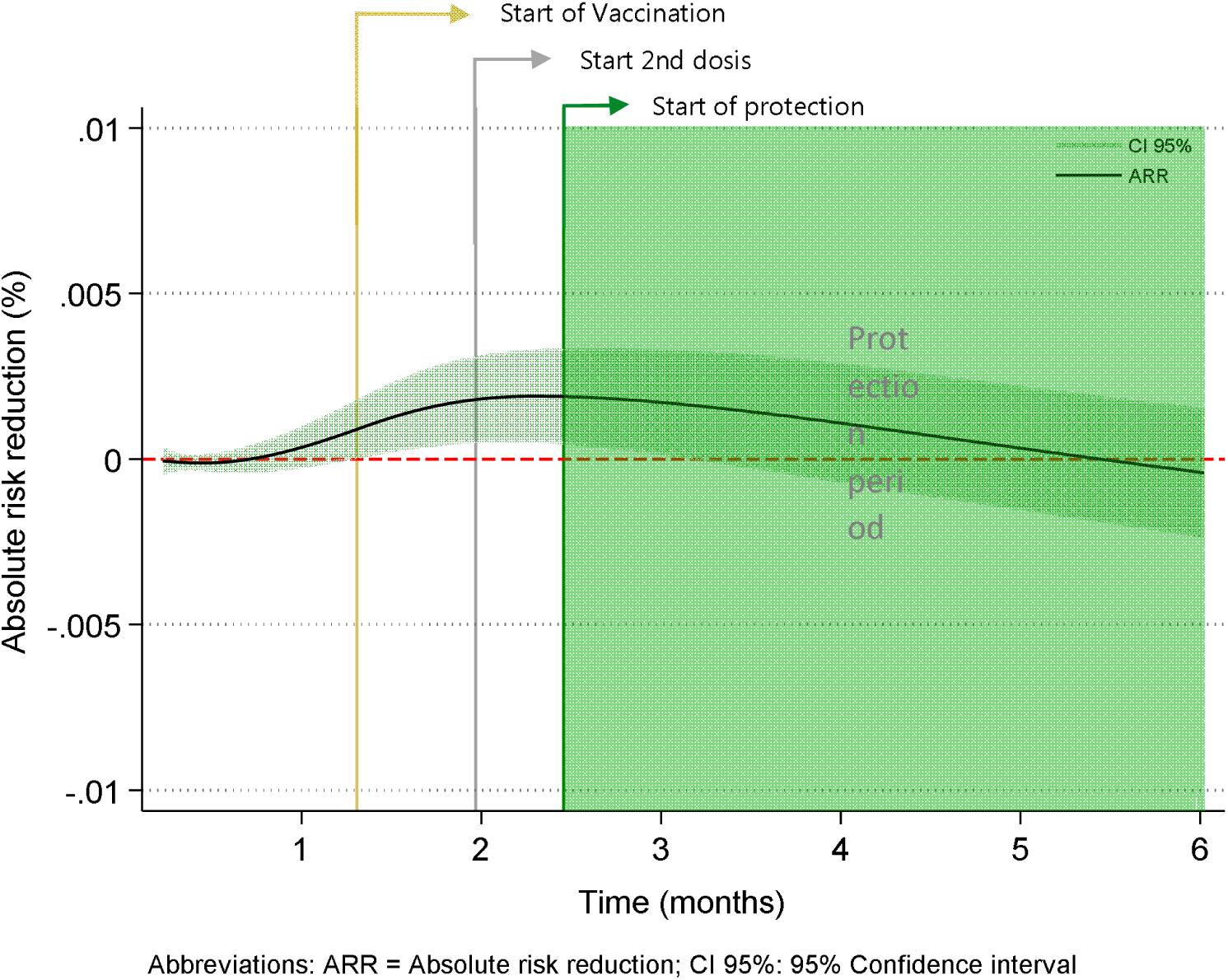
Dynamics of the risk of death ratio (ARR) of health workers infected by SARS-CoV-2, between the first and second waves of COVID-19 in Peru, 2020-2021. (N = 32,159)

The risk ratio (Hazard Ratio) showed a higher incidence of mortality in the second wave compared to the first. The risk of dying of health workers in the second wave was twice the risk of dying in the first wave (HR = 2), before the start of vaccination. At the beginning of the vaccination protection period, the risk of dying in the second wave is reduced until it becomes like the risk of dying in the first wave (HR = 1). Finally, six months after the second wave and after three and a half times the start of vaccination protection, the risk of dying in the second wave is 87.5% lower compared to the first wave (HR = 0.125) (see graph 4).

**Graph 4.**
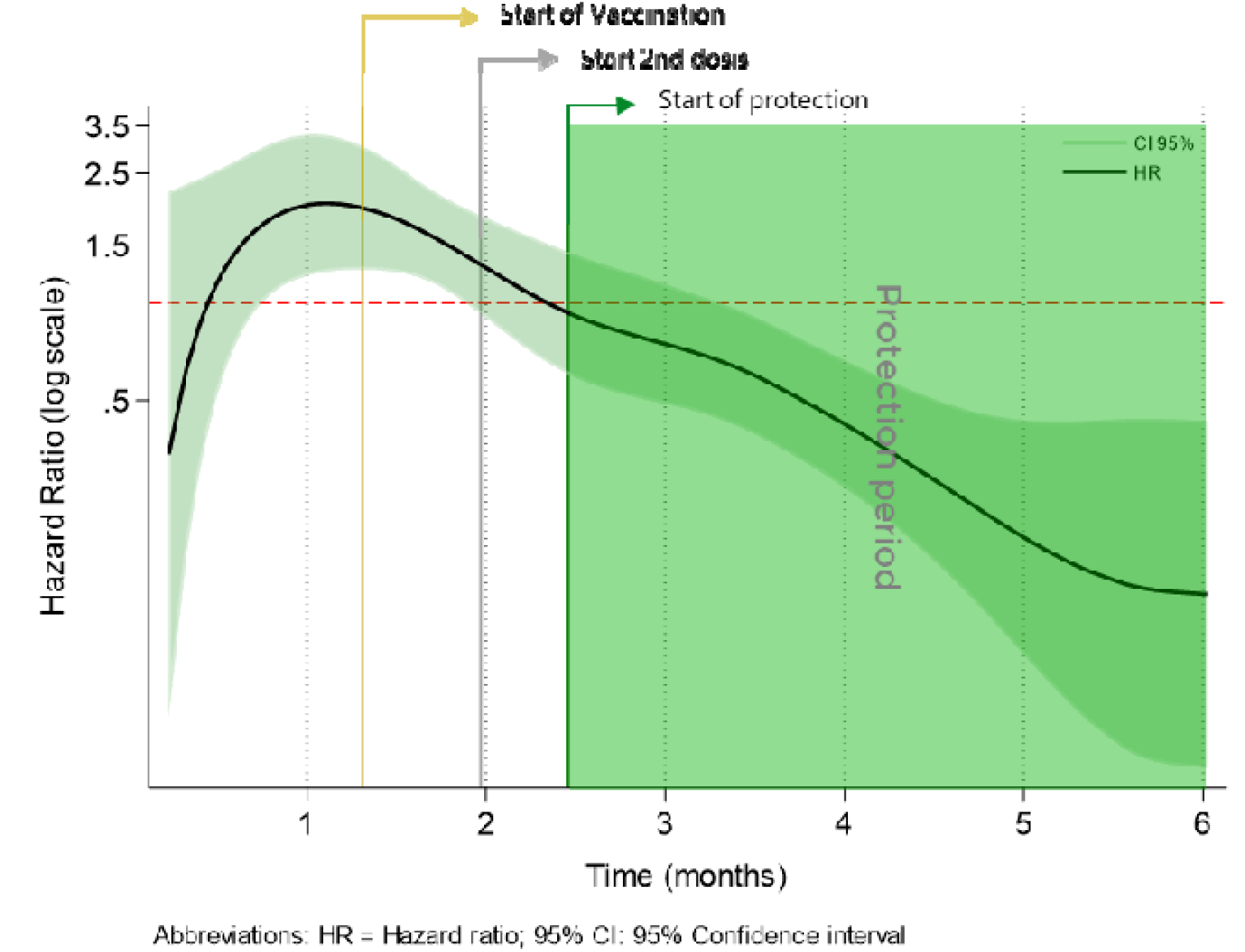
Dynamics of the death risk ratio (Hazard Ratio) of health workers infected by SARS-CoV-2, between the first and second wave of COVID-19 in Peru, 2020-2021. (N = 32,159)

When comparing the mortality curves of health workers during the first wave and during the second wave, by sex, a flattening of the survival curve is also verified during the second wave, from the beginning of the protection period. However, this is much more evident among male health workers (See graphs 5 and 6).

**Graph 5.**
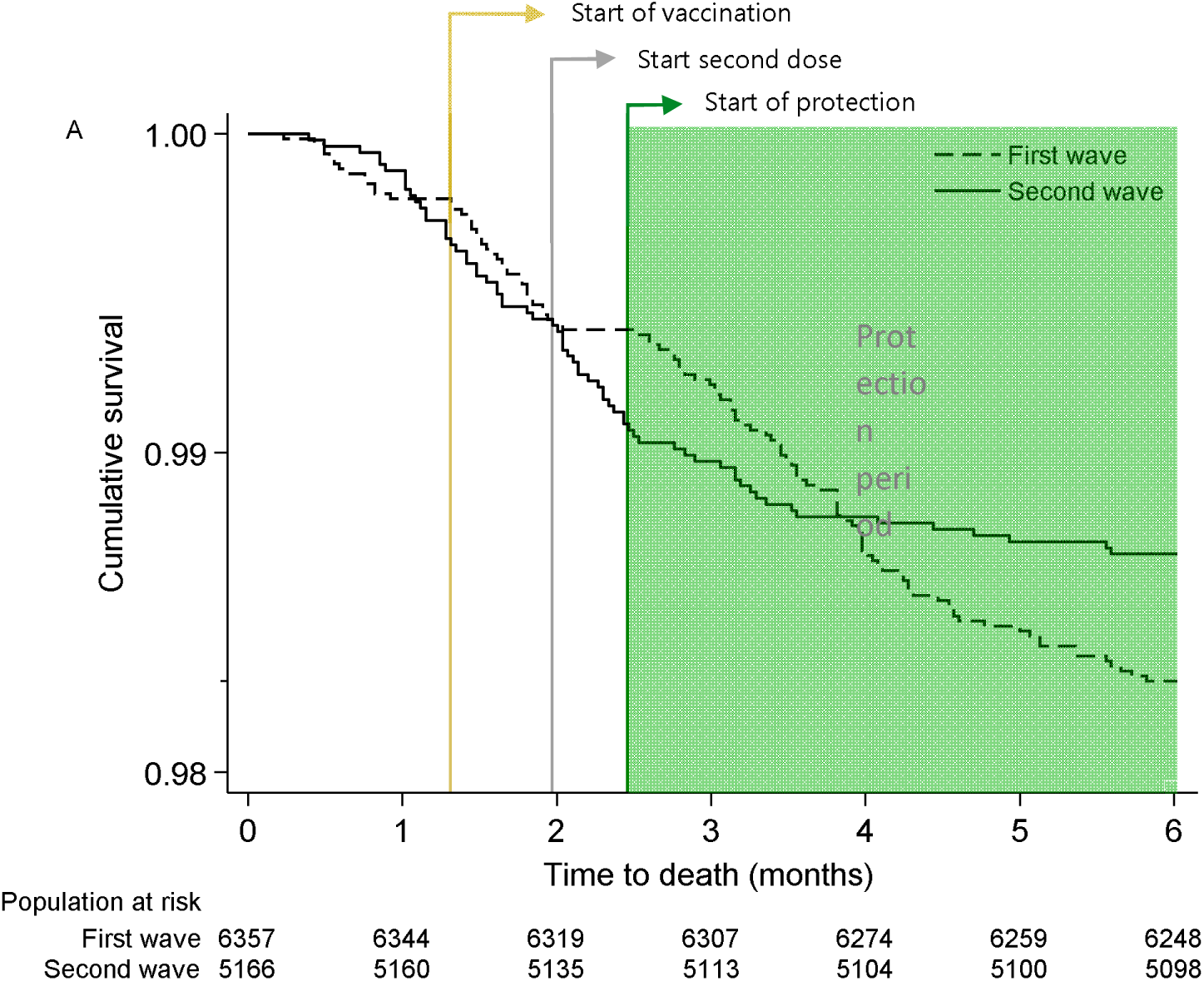
Survival levels (Kaplan Meier) of male health workers infected by SARS-CoV-2, during the first and second wave of COVID-19 in Peru, 2020-2021 (N = 11,523).

**Figure 6.**
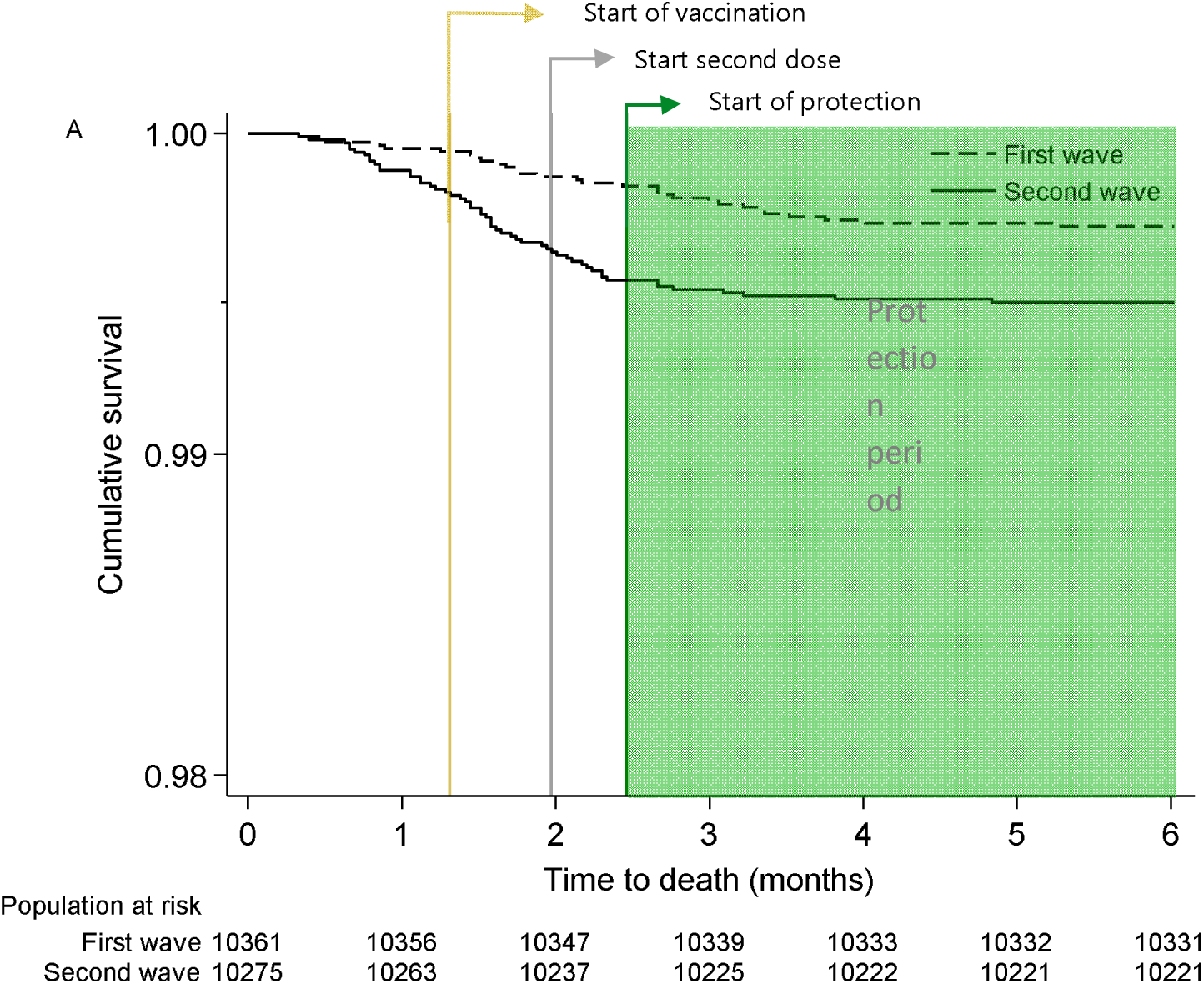
Graph 6. Survival levels (Kaplan Meier) of female health workers infected by SARS-CoV-2, during the first and second wave of COVID-19 in Peru, 2020-2021 (N = 20,636).

## DISCUSSION

The results of the present study show in a preliminary way the positive change in the survival of adult health workers in the context of the first phase of vaccination against COVID-19 in Peru. These results consistently showed that, after the vaccination period, the population of health workers improves its survival compared to the survival of the general population that is not health workers; and that health workers improves their survival during the second wave of COVID19 transmission in 2021, compared to their survival during the first wave in 2020.

In the first place, a higher level of survival of health workers was observed compared to the general population infected by SARS-CoV-2 during the first half of 2021. This situation could initially be explained by greater access to health care, higher socioeconomic level, and greater knowledge about the care regarding their illness of the health workers compared to the general population. However, the observed increase in the difference in survival between both groups increases after the period that followed the vaccination of health workers, which can be attributed to the effect of this intervention on the mortality of health workers.

Additionally, an improvement in the survival of health workers infected by SARS-CoV-2 was observed in the second wave, during the vaccination period in Peru, compared to the first. In this context, the higher mortality of health workers observed at the beginning of the second wave would be related to the higher mortality from COVID-19 that occurred at the national level. On the other hand, the comparison of risks through indicators (RAR, HR) showed that the initial increase in mortality of the second wave suffered a deceleration from the start of vaccination, turning towards a frank decrease in the period of protection, managing to cancel the difference in mortality between second and first wave. This suggests a possible impact of vaccination on mortality that would occur from the beginning of its application and would consolidate after the application of the second vaccination dose.

Finally, in the sub analysis carried out by sex, it was evidenced that male health workers infected by SARS-CoV-2 showed a greater change in the level of survival after the vaccination period, achieving not only canceling the difference in mortality risk of the second wave compared to the first, but surpassing it in the end of the study period. This situation was less evident in females due to the lower overall mortality from COVID-19 of this subgroup (6).

The present study has some limitations. In the first place, since we do not have nominal data on people vaccinated at the national level, it cannot be determined whether all the health workers included were finally vaccinated during the study period. However, given the wide vaccination coverage in Peru for this population, it is estimated that the percentage of unvaccinated health workers would be low, and their exclusion would not significantly modify the results found. Additionally, in comparing survival between COVID-19 waves, the use of retrospective data does not allow an accurate assessment of the 60-day prognosis of patients infected in the last weeks of the year study period. However, this bias would act by attenuating the differences in survival found between the first and second waves, which could be even greater if patient mortality data were included in the following two months. Another aspect in this analysis is the difference in the availability of Peruvian health resources to face the COVID-19 pandemic in the first and second waves. Although this situation has been able to favor the reduction of mortality in the second wave, the change in survival seen during the start of the first and second doses and the period of protection strongly suggest the contribution of the vaccination process in the change in survival of health workers.

The results found coincide with the evidence presented at the national level that suggests a potential influence of vaccination on the mortality rate in medical personnel, given the decrease in this indicator during the first months of the vaccination period in Peru (5). In the same way, this coincides with the evidence shown in countries such as Chile, the United States and the United Kingdom that have shown a marked reduction in mortality at a general level as the development of vaccination against COVID-19 advances in their respective countries (7) (8) (9).

In conclusion, a positive change is shown in the level of survival of health workers during the vaccination process against COVID-19 in Peru, especially in male personnel. This result, added to the evidence found at the national and international level, reaffirms the value of vaccination as one of the main strategies to combat the effects of the COVID-19 pandemic, especially in highly population exposed to the virus such as health workers.

Following this line, it is recommended to carry out future studies aimed at estimating the impact of vaccination against COVID-19 in Peru on the mortality of health workers in terms of effectiveness, as well as the evaluation of other indicators of interest such as infection, hospitalization, and admission to intensive care, which make it possible to assess its usefulness more precisely, as has been done in other countries (10) (11) (12).

## Data Availability

The data in this manuscript correspond to data collected by the Ministry of Health and by the National Institute of Health, as part of their competencies in the care of the COVID-19 pandemic, and are available with the respective authorizations.

